# DNA metabarcoding captures dietary plant diversity in individuals and cohorts

**DOI:** 10.1101/2022.06.13.22276343

**Authors:** Brianna L. Petrone, Ammara Aqeel, Sharon Jiang, Heather K. Durand, Eric P. Dallow, Jessica R. McCann, Holly K. Dressman, Zhengzheng Hu, Christine B. Tenekjian, William S. Yancy, Patrick C. Seed, John F. Rawls, Sarah C. Armstrong, June Stevens, Lawrence A. David

## Abstract

Eating a varied diet is a central tenet of good nutrition. Here, we develop the first molecular tool to quantify human dietary plant diversity by applying DNA metabarcoding with the chloroplast *trnL*-P6 marker to 1,001 fecal samples from 310 participants across four cohorts. The number of plant taxa per sample (plant metabarcoding richness, or pMR) correlated with recorded intakes in interventional diets (ρ=0.31) and with indices calculated from a food-frequency questionnaire in freely-chosen diets (ρ=0.40-0.63). In adolescents unable to collect validated dietary survey data, *trnL* metabarcoding detected 111 plant taxa, with 86 consumed by more than one individual and four (wheat, chocolate, corn, and potato family) consumed by >70% of individuals. Adolescent pMR was associated with age and income, replicating prior epidemiologic findings. Overall, *trnL* metabarcoding promises an objective and accurate measure of the number and types of plants consumed that is applicable to diverse human populations.

## Introduction

The recommendation to “eat a variety of foods” is a well-known component of public health nutrition guidance in the United States^1^. This advice reflects the fact that foods vary in their macro- and micronutrient content, and diets that include a range of foods are more likely to be nutritionally adequate. Empirical support for this recommendation was first produced in the early 1990s, when studies conducted in the US and globally demonstrated that counts of individual foods or food groups could serve as proxies for nutrient adequacy^2–4^ or overall health^5^; these findings prompted the introduction of dietary diversity-specific measurement tools by the Food and Agriculture Organization^6^. Since the 2000s, diversity metrics have continued to be applied in studies of the relationship between overall dietary patterns and health outcomes^7–9^.

However, a unified framework for the impact of dietary diversity on health has been hampered by the enormous range of species included in human diets, the number of possible diversity metrics, and weaknesses in available dietary assessment tools. Considering plant foods alone, lists of consumed species range from the hundreds (354 of commercial importance^10^; 866 crops^11^) to the thousands (4,079 compiled from published lists^12^) out of tens of thousands of theoretically edible wild plants. To make this complexity manageable, most existing dietary diversity assessments query a small number of locally important foods or aggregated food groups. This selection can be done in many ways, leading to a proliferation of disparate tools: for example, the Dietary Diversity Score (DDS) has 6-, 9-, 13-, and 21-food group formulations^4^; the Food Variety Score (FVS) totals items from a restricted 45-item list^2^; and species richness (SR) enumerates unique plant, animal, or fungal sources of consumed foods^13^. Diversity scores rely on self-reported dietary data, which have well-characterized random and systematic errors^14^: individuals can forget roughly 20% of the foods they eat when completing diet recalls^15^, survey tools incompletely capture ethnic or minority foods^16^, and social desirability bias leads to underreporting of intake^17^. Self-reported meals must be disaggregated into ingredients and manually mapped onto selected food groups or species of origin by researchers, which often requires local expertise or direct observation of cooking and eating^6^. Together, these challenges emphasize a need for objective, standardized diversity assessment tools that collect data that can be readily compared across studies^3^.

Objective dietary biomarkers are alternatives to or validation for survey-based dietary diversity assessment. Biomarkers have been developed for total energy intakes^18^, individual nutrients (*e*.*g*. protein^18^), and food groups (*e*.*g*. whole grains^19^). However, to our knowledge no biomarkers for dietary diversity exist. “Omics”-scale methods are well-suited for this type of comprehensive dietary assessment^20,21^ but have not been developed for dietary diversity because many metabolite or protein signals do not uniquely identify food items or are transformed by the body prior to detection. In contrast, DNA sequencing methods can be applied to non-invasively collected samples from the digestive tract (*i*.*e*., stool) to identify food items with specificity that ranges from the phylogenetic family to species level. Relying on the fact that nearly all foods are derived from once-living organisms whose tissues contain DNA, variable regions in food genomes serve as molecular “barcodes” that can be amplified and sequenced from stool. Such “DNA metabarcoding” methods have been widely applied to study the diets of wild animals^22,23^ and in proof-of-concept work to human stool^24^ and stomach contents^25^. However, they have not been applied at scale in the setting of nutrition research, nor have they been used to calculate within-sample diversity, a practice commonplace in ecosystem and animal studies (where within-sample diversity is termed “alpha diversity” and species counts are termed “richness”)^26^.

To meet the need for a standardized biomarker for dietary diversity, we develop here DNA metabarcoding with the chloroplast *trnL*-P6 marker^27^ as a tool for dietary plant diversity assessment in humans and measure its performance in four distinct cohorts. We first report several protocol adaptations that make *trnL* metabarcoding more reliable in human samples. Next, we test the relationship between *trnL* metabarcoding-based plant diversity and (1) recorded plant diversity of interventional diets and (2) indirect measures of dietary diversity from validated dietary self-reports. Finally, we transition to proof-of-concept testing to show that *trnL* metabarcoding measures are a valuable tool for testing epidemiological hypotheses and exploratory analysis of cohorts with limited dietary data.

## Results

### *trnL* METABARCODING PROTOCOL DEVELOPMENT

We developed a molecular approach for measuring dietary plant diversity by amplifying and sequencing residual DNA from human stool samples using the *trnL*-P6 region of the chloroplast genome (“*trnL* metabarcoding”). To accurately measure plant metabarcoding richness (pMR, the number of plant taxa detected per sample with *trnL* metabarcoding; **Fig. 1a**), we followed recommendations for microbiome metabarcoding studies^28^ to refine to our prior *trnL* metabarcoding protocol^24^, which was capable of detecting dietary plant taxa but limited by a low PCR success rate (∼50%). We moved from a standard- to a high-fidelity polymerase to reduce PCR errors and facilitate accurate taxonomic assignment of *trnL* sequence variants, and switched from a one-step to a two-step amplification protocol to avoid bias from barcode differences in the primary amplification and reduce primer synthesis costs. In the primary amplification, we adjusted reaction annealing temperature from 55 to 63°C to reduce formation of non- specific products and maximize amplification and sequencing yields (**Fig. S1a-c**), which further improved when template volume was quadrupled (**Fig. S1d-f**). In the second amplification step, an increase from 8 to 10 barcoding PCR cycles improved yields a further 3.4-fold. In samples tested pre- and post- optimization, these changes collectively resulted in median increases of 2.2 ng/μl in amplified DNA, 18,600 *trnL* sequencing reads, and 4 additional plant taxa detected per sample (*n*=199, **Fig. S1g-i**). Overall, the optimized protocol had a PCR success rate of 92%.

**Figure 1.**
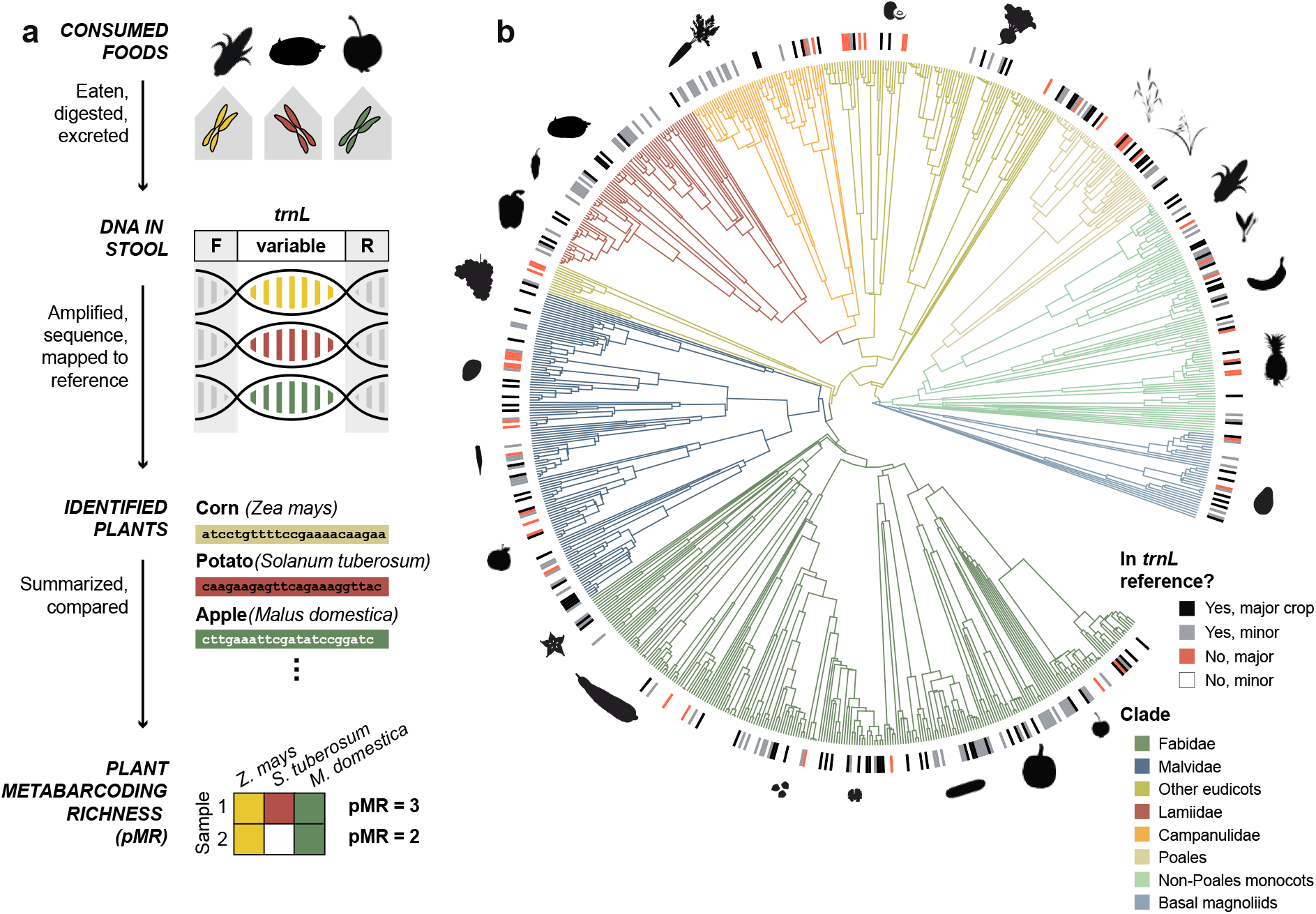
Calculation and dietary scope of plant metabarcoding richness (pMR). **(a)** Conceptual overview of *trnL* metabarcoding protocol and pMR calculation. Conserved primers (F, R) flank a variable *trnL* region, allowing amplification of a mixed pool of plant food-derived DNA from stool. **(b)** The reference *trnL* sequence database had broad representation (black and gray tick marks in outer ring) of food crop species^11^ (full phylogenetic tree) and included multiple sequences for 27% of plant taxa, which indicates within-food genetic variation at the *trnL*- P6 locus. Leaves in the crop tree terminate at the species level, though 70 subspecies- and 52 variety-level taxa were included in the full reference. Major plant crops were more likely to be included in the reference (Chi-square 188.94, df = 2, p < 10^−15^). Example plants from each clade are shown in silhouette. Clockwise from legend, these are: apple, pumpkin, cucumber, walnut, chickpea, cassava, starfruit, orange, okra, mango, grape, bell pepper, chili pepper, potato, carrot, kiwi, beet, rice, wheat, corn, onion, banana, pineapple, and avocado.

Bioinformatically, our optimization focused on assigning *trnL* sequencing reads to a more extensive reference database. In our previous work, 27% of reads did not have an exact match to a sequence in the reference database and could not be included in subsequent analyses. We therefore expanded our reference database of dietary plants from 185 sequences (representing 72 species) to 853 sequences (470 species covering 62% of all families and 83% of major crop families from a recent food plant phylogeny^11^, **Fig. 1b**). To reduce the percentage of unassigned reads and improve dietary diversity estimates, we shifted from grouping similar DNA sequences into operational taxonomic units to inferring exact amplicon sequence variants (ASVs), which enabled diversity estimation independent of a reference assignment and straightforward merging of metabarcoding datasets from multiple sequencing runs^29^. In concert, our expanded reference database and new pipeline reduced the percentage of unmapped reads to 0.9% (per-sample median 0.2% and median absolute deviation 0.3%; **Fig. S2**).

### *trnL* METABARCODING VALIDATION FOR INTERVENTIONAL DIETS

With our optimized experimental and bioinformatic protocols, we examined the potential for *trnL* metabarcoding to capture recorded dietary plant diversity using a cohort of individuals undergoing a targeted dietary intervention (“Weight Loss”; **Table 1**; *n=*41 samples from 4 individuals). Members of the Weight Loss cohort were clients of a residential-style, medically supervised weight loss center, with all their weekday meals prepared in the center’s cafeteria and consumed on site. Meal recipes and participant orders were logged by a digital menu system, which allowed us to specify a plant taxon for 96% of the 425 unique plant-derived food items consumed, including those from complex meals (*e*.*g*., “Mushroom Wild Rice Pilaf” could be separated into wild rice, white rice, portobello mushrooms, onion, pecans, thyme, parsley, and sage).

Median pMR ranged from 13.5 to 26 taxa in Weight Loss participants, which encompassed the median range of 22 to 26 plants per individual per day expected from diet records. On a per-sample basis, pMR was positively correlated with the number of food species calculated from coded menu data on the day prior to stool collection (Spearman ρ=0.31, *p*=0.04, **Fig. 2a**), and unrelated to tomorrow’s menu or a randomly selected menu day (**Fig. S3a**,**b**). pMR estimates of dietary plant diversity were slightly higher than menu data, with an average error of 4 taxa (average absolute error 7 taxa) above the recorded value. This difference could indicate that pMR may measure >1 day of food intake (*i*.*e*. if gastrointestinal transit time is >24h) or report food not captured by digital menus, including a daily fruit offering, selections from the cafeteria salad bar, or off-menu eating that occurred when clients were not at the center. Thus, in a small, controlled setting with high-quality dietary data available for comparison, our data indicated that pMR measured from stool was related to recorded dietary plant diversity.

**Figure 2.**
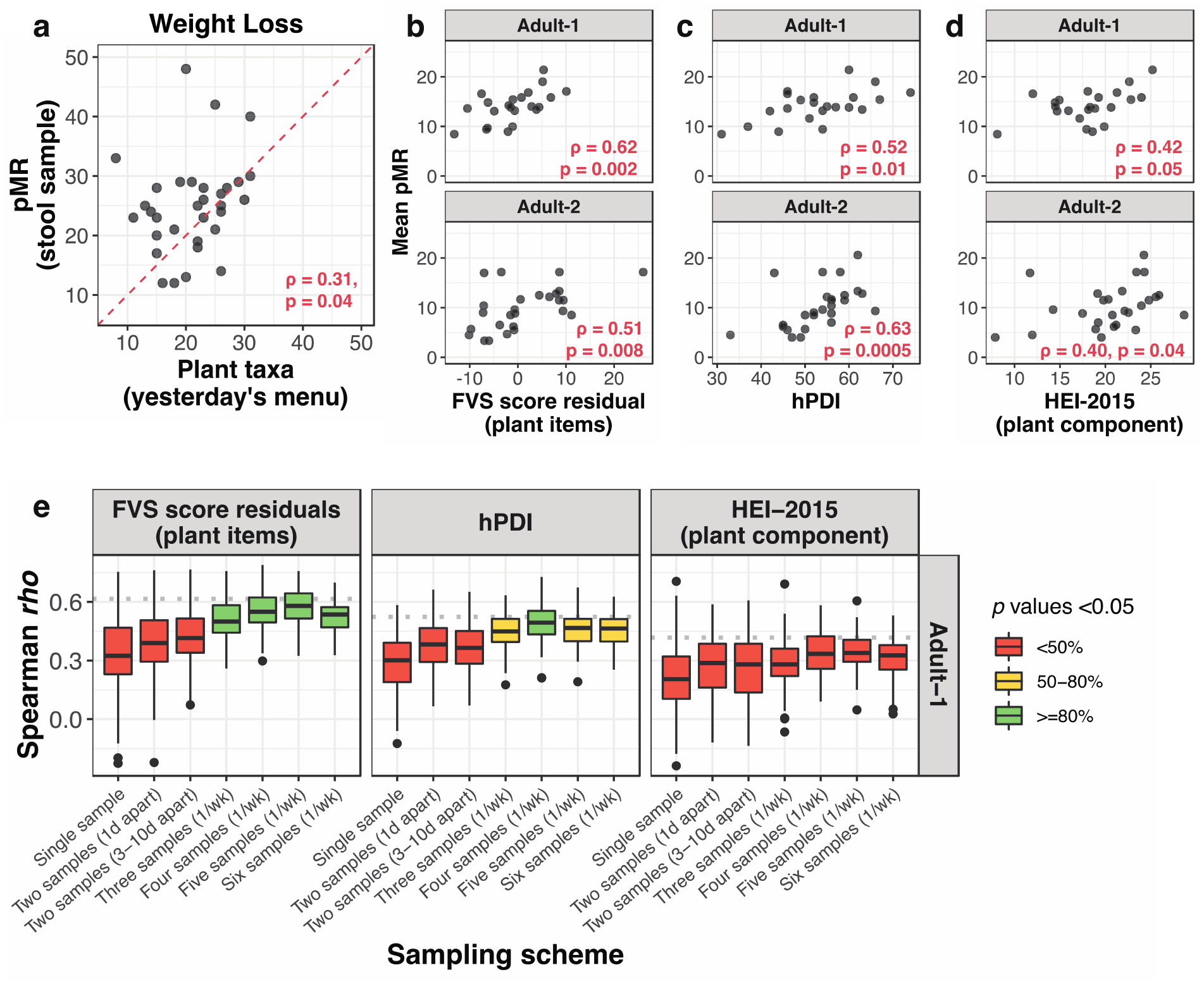
pMR is associated with independent measures of dietary diversity and quality. **(a)** Correlation between pMR and number of plant taxa from recorded menus of Weight Loss participants from the day prior to stool collection. The red dotted line denotes a theoretical perfect correspondence between the two measures. **(b-d)** Correlations between mean pMR (pMR averaged across all available stool samples per participant) and dietary diversity **(b)** and quality **(c, d)** indices derived from FFQ data in Adult-1 and Adult-2 participants. **(e)** Correlations from upper panels of **(b-d)** re-tested under candidate sampling schemes with mean pMR derived from a smaller number of stool samples. The “two samples (3-10d apart)” is the current dietary assessment protocol used by the National Health and Nutrition Examination Survey (NHANES)^38^. All boxplots represent ∼100 random subsamples at each strategy, and color indicates the percentage of iterations reaching the statistical significance threshold of p<0.05. Spearman correlations are one-tailed in **(a)**, and two tailed in **(b-e)**.

### *trnL* METABARCODING VALIDATION FOR FREELY-CHOSEN DIETS

Because the Weight Loss cohort consumed an interventional health-promoting diet and dietary diversity measurement is of importance across the real-world spectrum of intakes, we next extended pMR to extracted fecal DNA available from two larger, free-eating adult cohorts that completed food-frequency questionnaires (FFQs), a standard dietary assessment tool in nutritional epidemiology (“Adult-1” and “Adult-2,” *n*=28 and *n*=32, respectively; **Table 1**). Both cohorts^30,31^ were recruited for studies testing the impact of fiber supplementation on the gut microbiota. Participants ate their typical diets and collected multiple stool samples per week over a 6- or 3-week period (median of 16 and 6 samples/person for Adult- 1 and Adult-2, respectively).

To assess the degree of alignment between pMR and FFQ data, we summarized reported foods into dietary indices. We included a dietary diversity score, which counted unique food items, and two dietary quality scores, which weighted food items or groups based on amount consumed and health benefit or harm. For a diversity index, we selected the Food Variety Score (FVS), a score with a twenty-year history^3^ that correlates with nutrient adequacy^2^ and reduced risk of coronary heart disease and all-cause mortality^32^. For quality scores, we evaluated the Healthy Eating Index 2015 (HEI), which indicates adherence to US dietary guidelines, and the healthy, unhealthy, and overall Plant-based Dietary Indices (hPDI/uPDI/PDI), which assess plant presence and quality in the diet. We chose HEI and hPDI/uPDI because they have been previously linked to reduced risk of chronic disease morbidity or mortality^33–36^ and demonstrated to correlate tightly (ρ>0.7) with predictions based on microbiome composition^37^, which, like pMR, is a stool-based molecular measurement. Because HEI and FVS both include animal components, we used only the portion of the score that referenced plants, and because FVS scores scaled linearly with dietary calories, we used energy-adjusted residuals in place of the raw score (see **Methods**).

We identified significant, positive correlations between pMR and FFQ-based dietary indices in both Adult-1 and Adult-2 cohorts. Mean pMR per participant was positively correlated specifically with the plant component residuals of the Food Variety Score (FVS) (**Fig. 2b**; Adult-1 Spearman ρ=0.62, *p*=0.002, Adult-2 ρ=0.51, *p*=0.008); with the healthful component of the PDI (hPDI) (**Fig. 2c**; Adult-1 ρ=0.52, *p*=0.01, Adult-2 ρ=0.63, *p*=0.0005); and with the plant-based component score of the Healthy Eating Index 2015 (HEI-2015) (**Fig. 2d**; Adult-1 ρ=0.42, *p*=0.05, Adult-2 ρ=0.40, *p*=0.04). Correlations were absent or negative when tested against animal-based or unhealthy component scores alone (**Fig. S4**). All results except that for HEI-2015 were robust to rarefaction, a statistical downsampling to estimate richness in samples of varying sequencing depth (see additional details in **Methods, Table S2**). These findings indicate that pMR, a molecular dietary diversity measure with distinct sources of error from self-report instruments, can nevertheless rank individuals in a significantly similar way to multiple validated diversity and quality indices based on FFQ data.

Given the dense stool sampling protocols of the Adult-1 and Adult-2 cohorts, we next sought to determine the minimum number of samples per participant necessary to capture a comprehensive view of dietary plant diversity. We generated collector’s curves, an ecological tool used to assess richness as a function of sampling effort, for each participant (**Fig. S6a**). Unlike the average pMR calculated above, collector’s curves provide a running tally of the number of unique plant taxa detected as samples from the same individual are successively pooled. Curves were well-fit by a logarithm function, indicating that cumulative pMR plateaus with sufficient sampling. Consistent with prior work in diet records, which detected a plateau in “food repertoire” after 10 to 15 days of recorded intake^39^, the early plateau phase was often reached by individuals with >15 samples in the Adult-1 cohort, but rarely in Adult-2 participants, who collected at most 6 stool samples.

Even though a dozen stool samples may be required to observe an individual’s total potential dietary diversity, we found that averaged pMR from fewer samples could still reproduce the significant associations with dietary indices described above. Subsampling each participant recapitulated the significant correlation with hPDI at least 80% of the time and with FVS at least 50% of the time under at least one reduced sampling strategy in both cohorts (100 iterations at each strategy, unless fewer unique combinations were possible; Adult-1 in **Fig. 2e**; Adult-2 in **Fig. S7** due to more limited subsampling). The relationship to the HEI-2015 plant component score was not robust to subsampling, likely because it measures adequate intake of only five highly summarized food categories (*e*.*g*. “total vegetables”) and thus is better approximated by average pMR derived from larger number of samples. These results indicate that pMR from as few as three samples per person approximates the ranking of individuals by both a traditional dietary diversity index (FVS) and a dietary quality index (hPDI).

### *trnL* METABARCODING IN SETTINGS WITHOUT AVAILABLE DIETARY DATA

We next applied DNA metabarcoding in a setting where traditional dietary assessment measures were not collected. In a pediatric study of gut microbiota in adolescents with and without obesity from racially, ethnically, and socioeconomically diverse backgrounds (“Adolescent,” *n*=246, 79% with BMI >95^th^ percentile, 53% Black, 18% Hispanic, and >40% with household income <$50,000/year; **Table 1**), dietary assessment was limited to a custom 7-question survey. Two lengthier assessments had been eliminated within the first ten enrolled participants as they proved too cumbersome for families to complete.

Because no data on specific food items were available for the Adolescent cohort, we leveraged the taxon-level food identifications of *trnL* metabarcoding to identify plants included in participant diets. Across the cohort, we detected 111 unique *trnL* sequence variants, which came from 45 plant families, 89 plant genera, and 84 plant species. The most frequently observed food items were wheat or rye (the detected *trnL* sequence variant being the same for both foods; 96% of participants), chocolate (88%), corn (87%), and members of the potato family (a *trnL* sequence variant shared by potato, tomatillo, tamarillo, goji berry, cutleaf groundcherry, and edible nightshades; 71% of participants). However, the vast majority of *trnL* sequence variants had low prevalence across the pool of subjects, indicating that a small set of foods were commonly consumed and many more were unique to the diets of only a handful of individuals (**Fig. S8a**). The types of plant foods detected in the Adolescent cohort did not differ widely from the Weight Loss, Adult-1, or Adult-2 cohorts (**Fig. S8b**), contributed to a wide pMR range across the cohort (median 12 plant taxa per sample, median absolute deviation 4.4; **Fig. 3a**), and the presence or absence of foods in the diet indicated a spectrum of intakes rather than a partitioning of distinct eating patterns (**Fig. S8c**).

**Figure 3.**
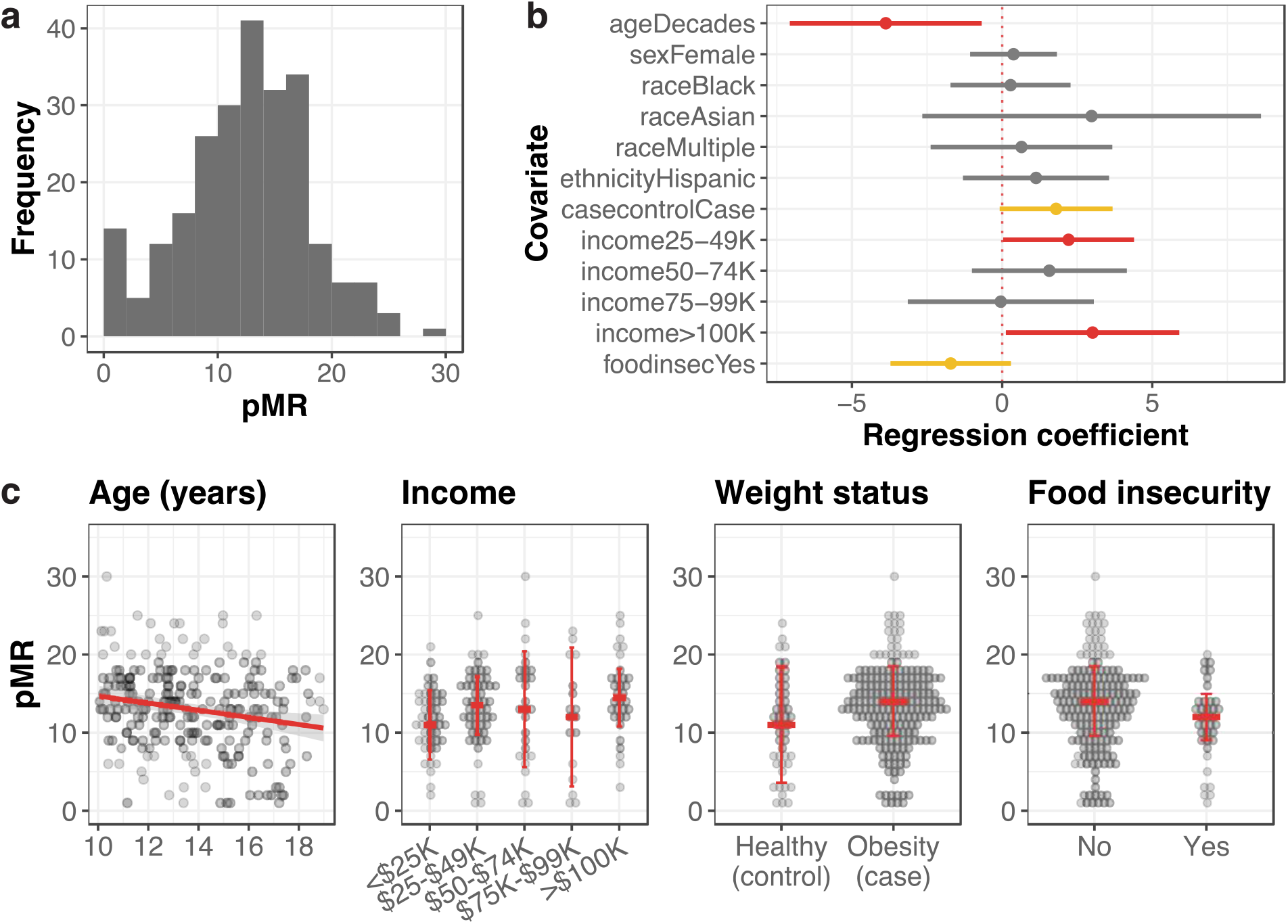
pMR detects known relationships between dietary diversity and demographic, health, and socioeconomic variables. **(a)** Histogram of pMR across Adolescent samples. **(b)** Visualization of linear model output, showing effect sizes and 95% confidence intervals of associations of demographic, clinical, and socioeconomic covariates with pMR as a response variable. Coefficient estimates with p≤0.05 are indicated in red, 0.05<p≤0.1 in yellow, and p>0.1 in gray. For categorical variables shown, the reference category is as follows: white for race, non-Hispanic ethnicity for ethnicity, control (healthy body weight) for case-control status, self-reported income <$25,000 annually for income, and no occurrence of food running out for food insecurity. Income and food insecurity both also included an “Unknown” category to accommodate missing responses, which we do not interpret and are not shown. **(c)** Raw data underlying significant or trend covariates from **(b)**.

The Adolescent cohort was more racially and ethnically diverse than the Weight Loss, Adult-1, and Adult-2 cohorts, and this enabled us to explore whether demographic variables were associated with pMR. In a multiple regression, pMR was negatively associated with age (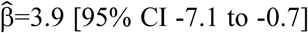, p=0.02), positively associated with higher income categories (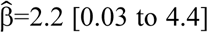, p=0.05 for $25,000-49,000/year and 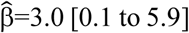, p=0.04 for $100,000/year, both relative to lowest bracket of <$25,000/year), trended higher with obesity status (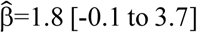, p=0.06), and lower with food insecurity (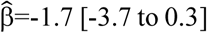, p = 0.09), and was unrelated to sex, race, and ethnicity (all with p>0.28; fitted coefficients in **Fig. 3b**, raw relationships in **Fig. 3c**). The negative association between pMR and age is consistent with data that American adolescents are less likely to eat dinner with their families as they age, which is associated with lower intakes of fruit, vegetable, and whole grain foods^40^. The trend between pMR and obesity status supports a recent recommendation to reduce emphasis on dietary diversity in adult populations^8^ given inconsistent associations with lower adiposity^41,42^ and positive relationships to total energy intake^43,44^, obesity^32,43^, and body fat percentage^44^ detected in some studies. Finally, the positive association between pMR and higher income categories aligns with epidemiologic studies within and outside the United States, which report increased dietary diversity in households with higher socioeconomic status^2,45–47^. We performed a comparable literature review for covariates for which we did not detect a significant association and found both concordant (ethnicity^48^) and discordant (sex^5,32^, race^32,46^) results, although all were derived from non-adolescent cohorts. Thus, dietary plant diversity measured by pMR recapitulates the majority of known epidemiological findings from studies that used self-report-based diversity measures.

## Discussion

In this study, we establish that dietary plant diversity assessment with *trnL* metabarcoding, a sequencing technique applied to non-invasively collected biological specimens, reliably reports the number of plant foods consumed by individuals. pMR as determined by *trnL* metabarcoding aligns well with diet records, validated survey tools, and demographic variables. This work, in which all but 4 of the 310 participants were consuming their typical diets, is the first large-scale application of *trnL* metabarcoding to free-eating individuals, who are difficult to reliably survey with existing dietary assessment instruments.

In total, we detected 187 unique *trnL* sequence variants representing 146 taxa, including plants from 73% of major food crop families present in the reference database. This expands from two prior *trnL* metabarcoding studies conducted in humans that detected 47 unique plant taxa across 11 individuals’ fecal samples^24^ and 124 across 48 individuals’ stomach contents^25^. The strength of positive correlations (Spearman ρ∼0.3-0.6) between pMR and indices calculated from validated survey tools falls within a range that is a workable proxy in dietary research: for example, the use of a simplified survey specifically for dietary diversity measurement is supported by Pearson correlations to nutrient adequacy from a more complex tool ranging from *r*=0.3-0.6^2,4,49^. Further, our findings suggest that pMR combines elements of existing dietary diversity and quality indices, which may indicate that food features like amount and quality are implicitly incorporated in pMR measurement (*i*.*e*., due to a detection bias in favor of items consumed in large quantities or without DNA degradation from industrial processing). This is useful given recent concern that strict definitions of dietary diversity do not discriminate against unhealthy eating: in studies of adults not at risk of nutrient inadequacy, increased dietary diversity associates with greater intake of processed foods, refined grains, and sweetened drinks^50^.

No dietary assessment technique is universal in scope and without limitations. pMR does not capture processing or cooking techniques used to prepare foods. Still, some existing dietary diversity measures do not consider food preparation (species richness^13^) or track prepared items without counting them towards the final score (*e*.*g*., “Oils and fats” and “Sweets” in the measurement of the Women’s Dietary Diversity Score^6^); as is, pMR correlates with measures of healthy eating that incorporate food preparation (**Fig. 2b**). Our definition of pMR as the number of plant food taxa per sample differs from conventional dietary diversity, which is calculated as number of foods or food groups over a reference period (usually 24h). Due to interindividual differences in gastrointestinal transit time, pMR could summarize food intake over a period from 24 hours to multiple days. Pairing *trnL* metabarcoding with transit time indicators (edible dyes or proxies like Bristol stool scale scores) to adjust for this effect may reveal even more robust associations between pMR and independent diversity metrics. As a strict measure of richness, pMR also leaves out much of the detail in raw metabarcoding data, including abundance and identity of each *trnL* sequence variant. Future work will focus on identifying foods that can be reliably quantified with *trnL* metabarcoding and implementing internal standards to make quantitative estimates more reliable^51,52^. Abundance data can then be used to calculate alternative diversity measures like dietary evenness and dissimilarity^8,9^. With 99.1% of reads in the data presented here assigned to a food taxon, we estimate the *trnL* reference database provides nearly complete coverage of foods consumed in Western diets, but its performance for global cohorts is likely lower and will be the target of future updates. Finally, we have characterized only plant diversity, omitting animal and fungal sources of dietary variation. As for *trnL*, animal and fungal DNA metabarcoding markers can be adapted to human diet to provide a comprehensive view of intakes.

Together, these findings position *trnL* metabarcoding as a candidate genetic biomarker of food intake, and support its use to derive pMR prospectively from any individual able to provide a stool sample and retrospectively from biorepositories. Unlike dietary surveys, *trnL* metabarcoding provides data in the conserved language of DNA sequence, which overcomes challenges of manual food item identification, grouping, and nomenclature^53^ and permits immediate harmonization of data across global studies. In practice, pMR can complement self-report tools by providing an alternative means of assessment in populations for whom validated dietary assessments do not exist or in settings where self-reports are limited by participant burden or resource constraints, as in the Adolescent cohort. Our *trnL* metabarcoding experimental protocols, bioinformatic pipeline, and reference database are publicly available and actively maintained (https://github.com/bpetrone/mb-pipeline). Estimated reagent costs per sample are $25 for laboratories with PCR capability and access to high-throughput sequencing technology and $85 for samples sent for sequencing at a core facility or commercial provider, competitive with diet survey costs (an interviewer-administered 24-hour recall costs $90 from a university-based provider). *trnL* metabarcoding therefore has the potential to be as widely available and accessible as 16S sequencing technology is for gut microbiome profiling from stool samples.

## Methods

### Study populations

Samples were drawn from three clinical trials and one sample biorepository, all based at Duke University in Durham, NC. The clinical trials consisted of a behavioral intervention that returned gut microbiome data to participants (NCT04037306, here “Weight Loss”) and studies assessing the impact of fiber supplementation on the gut microbiota^30^ (NCT03595306, “Adult-1”) and on human cognition, behavior, and physiology^31^ (NCT04055246, “Adult-2”). The sample biorepository was collected from adolescents with obesity and their healthy-weight siblings^54^ (NCT02959034, “Adolescent”). Application of *trnL* metabarcoding was a secondary analysis and determined exempt by the Duke Health Institutional Review Board (Pro00100567). Study characteristics and participant demographics are summarized in **Table 1**.

### Stool sample collection, processing, and DNA extraction

Stool samples were collected, stored, and DNA extracted as part of each primary study protocol. In all studies, stool was immediately frozen on collection by participants and transported frozen to a laboratory freezer. DNA extraction relied on versions of the PowerSoil kit system (QIAGEN, Hilden, Germany) following the manufacturer’s instructions. Briefly, Weight Loss, Adult-1, and Adult-2 samples were extracted with the DNeasy PowerSoil or MagAttract PowerSoil kit, depending on number of samples per processing batch; Adolescent samples were extracted with the DNeasy PowerSoil Pro kit. For Weight Loss, Adult-1, and Adult-2 samples, 1 to 1.5 grams of stool was slurried in PBS at a 10% weight-to-volume ratio in sterilized filter bags with a 0.33 mm pore size (Whirl-Pak, Madison, WI) using a Stomacher 80 Biomaster (Seward Limited, Worthing, United Kingdom). 750 μl was added to tube-based (PowerSoil) and 200 μl to plate-based (PowerSoil MagAttract) extractions. For Adolescent samples, whole stool was added directly to the extraction (PowerSoil Pro). Extracted DNA within each cohort was randomized and stored at -20°C prior to *trnL* metabarcoding.

### trnL *metabarcoding*

We performed *trnL* metabarcoding using a two-step PCR protocol. Primary PCR amplification of *trnL* used the KAPA HiFi HotStart PCR kit (KAPA Biosystems, Woburn, MA) in a 10 μl volume containing 0.5 μl of 10 μM forward and reverse primers (IDT, Coralville, Iowa), 2 μl of 5X KAPA HiFi buffer, 0.3 μl of 10 mM dNTPs, 0.1 μl of 100X SYBR Green I (Life Technologies, Carlsbad, CA), 0.1 μl KAPA HiFi polymerase, 3.5 μl nuclease free water, and 3 μl of extracted DNA template. The primers were *trnL*(UAA)*g* and *h*^27^ with Illumina overhang adapter sequences added at the 5′ end (**Table S1**). Cycling conditions were an initial denaturation at 95°C for 3 minutes, followed by 35 cycles of 98°C for 20 seconds, 63°C for 15 seconds, and 72° for 15 seconds. Each PCR batch included a positive and negative control, and samples were only advanced to the secondary PCR if controls performed as expected (otherwise, the entire batch was repeated). Secondary PCR amplification to add Illumina adapters and dual 8-bp indices for sample multiplexing was performed in a 50 μl volume containing 5 μl of 2.5 μM forward and reverse indexing primers (**Table S1**), 10 μl of 5X KAPA HiFi buffer, 1.5 μl of 10 mM dNTPs, 0.5 μl of 100X SYBR Green I, 0.5 μl KAPA HiFi polymerase, 22.5 μl nuclease free water, and 5 μl of primary PCR product diluted 1:100 in nuclease-free water.

### Sequencing library preparation

Amplicons were cleaned (Ampure XP, Beckman Coulter, Brea, CA), quantified (QuantIT dsDNA assay kit, Invitrogen, Waltham, MA), and combined in equimolar ratios to create a sequencing pool. If samples could not contribute enough DNA to fully balance the pool due to low post-PCR DNA concentration, they were added up to a set volume, typically 15-20 μl. Libraries were then concentrated, gel purified, quantified by both fluorimeter and qPCR, and spiked with 30% PhiX (Illumina, San Diego, CA) to mitigate low nucleotide diversity. Paired-end sequencing was carried out on an Illumina MiniSeq system according to the manufacturer’s instructions using a 300-cycle Mid, 150-cycle High, or 300-cycle High kit (Illumina, San Diego, CA, USA), depending on the number of samples in each pool.

### Reference database construction

A list of edible plant taxa was compiled from US food availability data^55^, global surveys^13^, and reference volumes^10^. DNA sequences likely to contain *trnL* were downloaded from two sources within NCBI: GenBank (all publicly available DNA sequence submissions) and the organelle genome resources of RefSeq (a curated, non-redundant subset of assembled chloroplast genomes). To obtain GenBank sequences, we used the entrez_search function of rentrez 1.2.3^56^ to submit separate queries for sequences containing “trnL” in any metadata field and each plant taxon name in the Organism field (*e*.*g*. “*Zea mays*[ORGN] AND *trnL*” to pull data for corn, or *Z. mays*). Sequences with an “UNVERIFIED:” flag were discarded. To obtain RefSeq sequences, the plastid sequence release current as of June 2021 was downloaded and subset to only those accessions including an edible taxon name. Results from either source were then filtered to sequences containing primer binding sites for *trnL*(UAA)*g* and *trnL*(UAA)*h* in the correct orientation. Binding sites were identified using a custom R script with a mismatch tolerance of 20% (≤3 mismatches for *trnL*(UAA)*g* and ≤4 for *trnL*(UAA)*h*), and sequence outside the primer binding sites removed. Identical *trnL* sequences from different accessions of the same taxon were de-duplicated, but we preserved distinct *trnL* sequences within taxa (indicating genetic variability) and identical *trnL* sequences from different taxa (indicating genetic conservation). The taxonomic tree of possible identifications in comparison to a plant food phylogeny (**Fig. 1b**) was visualized with ggtree v. 2.2.4^57^.

### Bioinformatic analysis

For each sequencing run, raw sequencing data were demultiplexed using bcl2fastq v2.20.0.422. Read-through into the Illumina adapter sequence at the 3’ end was detected and right-trimmed with BBDuk v. 38.38. Using cutadapt v. 3.4, paired reads were filtered to those beginning with the expected primer sequence (either *trnL*(UAA)*g* for the forward read or *trnL*(UAA)*h* for the reverse) and then trimmed of both 5’ and 3’ sequences using a linked adapter format with a 15% error tolerance. Reads were quality-filtered by discarding reads with >2 expected errors and truncated at the first base with a quality score ≤2. Amplicon sequence variants (ASVs) were inferred using DADA2 v 1.10.0. Taxonomic assignment was done with DADA2’s assignSpecies function, which identified ASVs by exact sequence matching to the custom *trnL* reference database, with multiple matches allowed. If multiple matches occurred, reads were assigned to the taxon representing the last common ancestor of all matched taxa (*e*.*g*. an ASV matching to both wheat [*Triticum aestivum*] and rye [*Secale cereale*] was relabeled as Poaceae, the family shared by both genera). Sequence data were screened for contamination on a per-PCR batch basis using decontam v.1.8.0 using DNA quantitation data from the library pooling step, and suspected contaminants were removed. ASV count tables, taxonomic assignments, and metadata were organized using phyloseq v. 1.32.0.

Prior to calculating plant metabarcoding richness (pMR), ASVs identified to the same food taxon at the species, subspecies, or variety level were merged to make pMR representative of food identity, rather than *trnL* sequence variation. ASVs representing distinct subsets of species in the same family or genus, which occur due to the last common ancestor method above, were identified and preserved as distinct (*e*.*g*. in the family Rosaceae, the rosids, one sequence variant indicates apple and pear intake and a second identifies strawberries and raspberries, so these were not merged). Plant metabarcoding richness (pMR) was then calculated as the number of unique taxa observed with at least one read count in each sample.

### Dietary data collection and processing

#### Digital menus (Weight Loss)

Complete menu data for each participant was exported from RealChoices menu software (SciMed Solutions, Durham, NC) and linked to ingredient names from recipe source files. Ingredient common names were then manually identified to plant species using the NCBI Taxonomy Browser and Integrated Taxonomic Information System databases. For ingredients that were themselves composite foods (*e*.*g*., “whole wheat bread”), we identified a primary ingredient using either provided brand information or the USDA FoodData Central database, which includes taxon mapping under the “Other information” header.

#### Dietary surveys (Adult-1 and Adult-2)

Habitual dietary intake over the past 1 month was assessed by administration of National Cancer Institute Diet History Questionnaire III (DHQ3), a 135-item, semi-quantitative food frequency questionnaire (FFQ). FFQ data were quality checked by estimating participant basal metabolic rate (BMR) using the Harris-Benedict equation^58^, calculating the ratio of reported calorie intake to estimated BMR, and excluding FFQs where this ratio was greater than two absolute deviations outside the median of the full dataset (corresponding to a ratio of <0.22 or >1.75) from further analysis, as done in a prior study^37^. This criterion preserved 87% of completed FFQs in the dataset (excluded responses were all for suspected underreporting).

##### Food variety score (FVS)

The Food Variety Score (FVS) was calculated as the number of unique food items consumed at least once per week. After summing daily intake frequencies within each food item, we tallied items with a daily frequency of consumption ≥0.14 (equivalent to 1/7, or a weekly frequency, as previously done for calculating FVS from frequency data^32^). The plant component of the overall FVS was calculated using the same procedure after manually labeling food items derived from plants or including a plant ingredient. Total and plant component FVS were then adjusted for overall calorie intakes using the nutrient residual method^59^: briefly, a linear regression model was used to fit FVS to overall energy intake in kilocalories, and the residuals from the model were used in place of raw FVS values.

##### Healthy Eating Index 2015 (HEI-2015)

The HEI-2015 and its component scores were calculated automatically by the DHQ3. We defined a plant HEI score as the sum of exclusively plant-based adequacy components (Total Vegetables, Greens and Beans, Total Fruits, Whole Fruits, and Whole Grains), which give higher scores to higher intakes of encouraged plant food groups. Conversely, we defined a non-plant-based HEI score as the sum of components with exclusively non-plant-based items (Dairy, Sodium, Added Sugars, and Saturated Fat). Saturated Fat may contain plant items like palm oil or coconut, but we expect this category is largely reflective of meat and dairy intake. Though meat and seafood are included in HEI component scores, their categories also include plant-based items (legumes for “Total Protein” and legumes, nuts, seeds, and soy for “Seafood and Plant Protein”). We therefore did not include these categories in either score definition above.

##### Plant-derived dietary index (PDI)

The PDI and its variations, healthy PDI (hPDI) and unhealthy PDI (uPDI), were calculated from DHQ3 data by manually assigning food items to specified food groups (*n*=18), splitting participants into quintiles based on gram weight of intake of each food group, and then scoring the quintiles from either 5 to 1 or 1 to 5, depending on the index being calculated. Food group scores were then summed within each participant to give the overall score. In rare cases, enough participants did not report consuming the food that they could not all be accommodated by the first quintile of the data; in this case, all participants with zero intake were assigned to the first quintile, and the remainder of the data split into quartiles and assigned to the 2^nd^ to 5^th^ intake categories.

### Statistical analysis

#### Weight loss cohort

One-tailed Spearman correlation between pMR and menu plant taxa was computed using the cor.test function from R stats v. 4.1.3. Average and absolute error were computed by subtracting the number of plant taxa recorded in the menu from the number detected by *trnL* metabarcoding and taking the mean or the absolute value of the difference, respectively. Samples collected on days following a weekend day (*i*.*e*. Sunday or Monday; *n* = 8) were excluded from the paired analysis because the on-site cafeteria only provided breakfast on weekends, and digital menus had to be supplemented with less accurate self-reports.

#### Adult-1 and Adult-2 cohorts

Two-tailed Spearman correlations were calculated between mean pMR (averaged across all samples for each participant) and FFQ data. For each subsampling scheme, samples that fit each strategy were randomly selected from the total available for each participant, and Spearman correlations were calculated using the mean pMR of only those samples. 100 subsampling iterations were performed for each scheme, unless fewer unique combinations were available or duplicate subsamples occurred by chance (this resulted in a loss of no more than three iterations from any combination of study, dietary index, and sampling scheme).

#### Adolescent cohort

Demographic, health, and socioeconomic status variables were included as covariates in a linear model with pMR as the outcome variable. All covariates were checked for completeness and missing entries coded as “Unknown” (*n=*62 for income, and *n*=28 for food insecurity) so as not to exclude missing data. We chose not to impute missing values because we hypothesized that missing responses to socioeconomic questions likely violated assumptions that data are missing completely at random (*i*.*e*., individuals in lower income or food-insecure categories would be more likely to leave the question blank). Because only 138 of 246 subjects (56%) had two timepoints, we used a linear model of pMR from the “Entry” timepoint alone rather than a mixed-effects model with repeated measurements. The distribution of pMR was approximately normal (tested with the descdist function of fitdistrplus v. 1.1.8), so we tested both a linear model using the lm function of R stats v. 4.1.3 and a negative binomial family generalized linear model (GLM) using the glm function, which as a discrete distribution is a theoretically better approximation of pMR. Both yielded similar results and we present the findings of the linear model here for simpler interpretation of the magnitude of fitted coefficients. We screened for, but did not detect, collinearity amongst model predictors using the function vif of car package 3.0.12. Observed versus predicted pMR and residual versus predicted pMR plots were generated to check model validity.

### Rarefaction

Rarefaction was performed using vegan v. 2.5.7 and statistical tests above repeated using rarefied pMR in place of raw pMR. Rarefaction provides a statistical estimate of richness that adjusts for variation in sequencing depth, which we first noted in the Adult-1 and Adult-2 cohorts (range 1-150,330, **Fig. S5a**) despite experimental strategies to balance samples within each sequencing batch. Because richness scales with sampling effort^26^ (**Fig. S5b**), we tested whether using rarefaction (statistical downsampling to a shared read depth) to adjust for differences in sequencing depth affected relationships between pMR and dietary data. Rarefaction strengthened the correlation between pMR and recorded menus in the Weight Loss cohort; in the Adult-1 and Adult-2 cohorts, rarefaction retained the positive correlations to FVS and hPDI at only slightly weakened magnitude, but rendered the relationship to HEI-2015 plant component score insignificant (**Table S2**). One interpretation of these findings is that read depth may indicate plant content of the diet rather than technical variation in sample preparation. In support of this hypothesis, FVS plant residuals, overall PDI, and HEI-plant component scores were all significantly lower for Adult-1 and Adult-2 samples with fewer than 1,000 reads, indicating reduced plant intake by an independent measure (**Fig. S5c)**. Therefore, we continued subsequent analyses without rarefaction (while monitoring its effects in **Table S2**).

## Data Availability

*trnL* metabarcoding data for all samples will be deposited to the European Nucleotide Archive prior to publication. De-identified clinical metadata associated with this study are available upon request and will be shared when consistent with applicable study agreements, regulations, and ethical standards.

## Data and code availability

*trnL* metabarcoding data for all samples will be deposited to the European Nucleotide Archive prior to publication. De-identified clinical metadata associated with this study are available upon request and will be shared when consistent with applicable study agreements, regulations, and ethical standards. R scripts to reproduce results from raw phyloseq objects are available on GitHub (https://github.com/bpetrone/plant-richness).

## Acknowledgements

We thank our study volunteers for their participation; Verónica Palacios for human study support; Michelle Kirtley for manuscript edits; Pao-Hwa Lin for guidance on dietary indices; and Tonya Snipes, Lisa Alston-Latta, and Margaret Huggins for keeping our lab spaces and glassware clean. This work was supported by the NIH (5R24DK110492-05 to JFR and PCS; 5R01DK116187-05 to LAD), the Burroughs Wellcome Fund Pathogenesis of Infectious Disease Award (LAD), a Duke Microbiome Center Development Grant (LAD), a Springer Nature Limited Global Grant for Gut Health (LAD), a fellowship from Integrative Bioinformatics for Investigating and Engineering Microbiomes (IBIEM) program (BLP), the Triangle Center for Evolutionary Medicine (BLP), and the Duke Medical Scientist Training program (BLP), and used a high-performance computing facility partially supported by grants 2016-IDG-1013 (“HARDAC+: Reproducible HPC for Next-generation Genomics”) and 2020-IIG-2109 (“HARDAC-M: Enabling memory-intensive computation for genomics”) from the North Carolina Biotechnology Center.

## Author contributions

BLP, SJ, and HK Durand performed *trnL* metabarcoding and conducted experimental optimization. BLP, HK Durand, LAD, CBT, and WSY coordinated the Weight Loss study; BLP and EPD received and processed Weight Loss participant samples. SJ coded Weight Loss participant menu data. HK Durand, SJ, and EPD processed Adult-1 and Adult-2 samples. JRM, PCS, JFR, and SCA coordinated the Adolescent study; JRM, HK Dressman, and ZH processed Adolescent samples; JRM curated Adolescent clinical and demographic data; and AA and BLP analyzed data from the Adolescent cohort. BLP analyzed all other data. BLP and LAD conceptualized the study and wrote the manuscript and JS provided guidance and feedback. All authors read and approved the manuscript.

## Extended data

**Figure S1.**
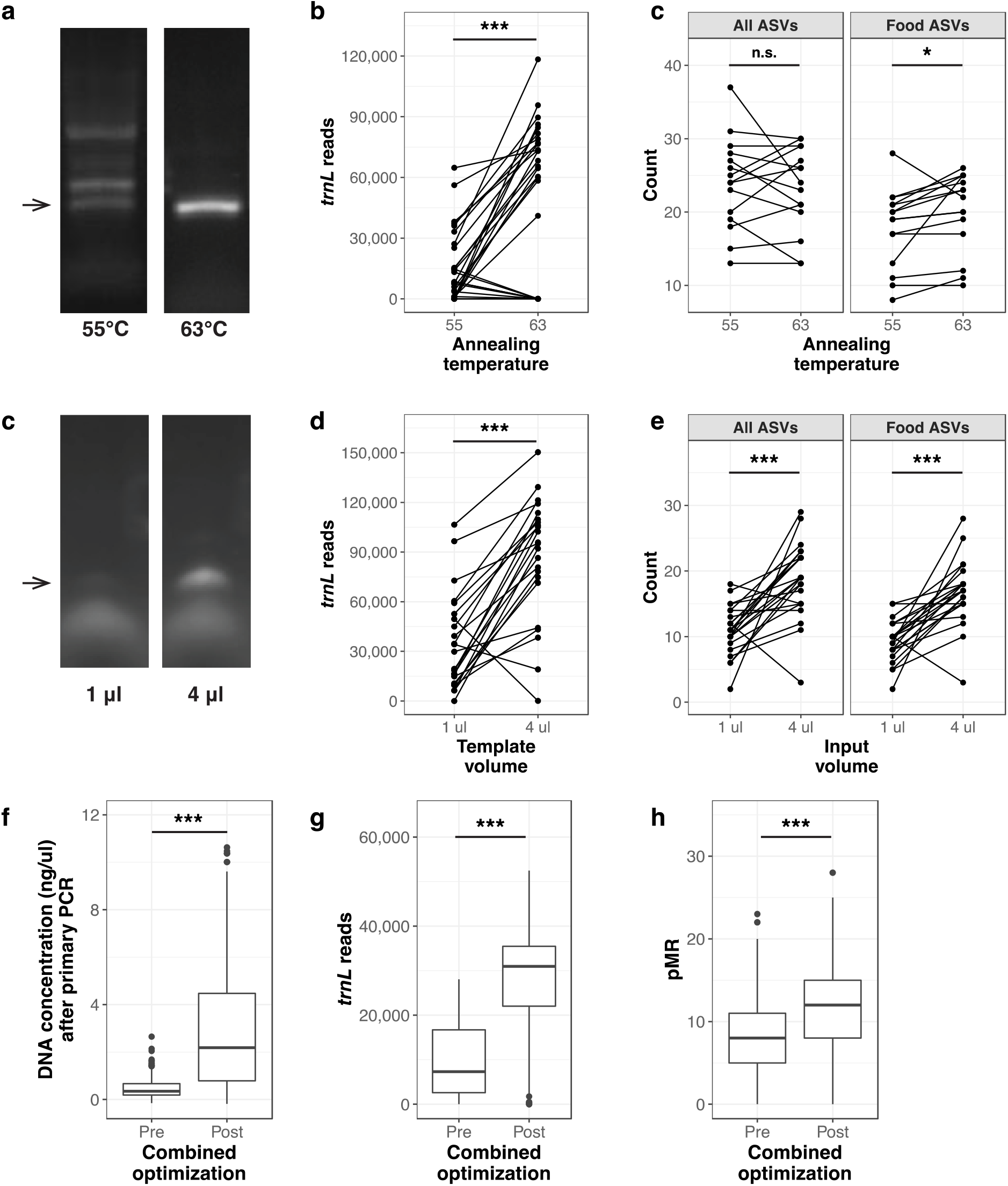
Experimental optimization of a prior trnL metabarcoding protocol improves dietary DNA amplification and sequencing. **(a)** Non-specific product formation (>143 bp, the upper limit of *trnL* length, indicated by arrow) is reduced at higher annealing temperatures. **(b)** Samples amplified with a 63°C annealing temperature have higher *trnL* sequencing read depth. Lines connect identical samples tested under either condition (n=28). **(c)** More food ASVs (identified by the *trnL* reference) were detected in samples amplified at 63°C despite no significant difference in total ASVs, indicating a reduction in non-specific amplification (n=15). **(c)** *trnL* PCR product formation (arrow) is increased with addition of more template volume to the reaction. **(d)** Samples with higher template volume added to the primary PCR have higher *trnL* sequencing read depth (n=23). **(e)** Higher template volume in the primary PCR increases the number of detected *trnL* ASVs (n=23). **(f-h)** Collectively, the suite of protocol changes improved **(f)** DNA concentrations after the primary amplification, **(g)** *trnL* reads by sequencing, and **(h)** the number of *trnL* ASVs per sample (n=199 samples evaluated pre- and post-optimization). All statistical tests shown are paired-sample Wilcoxon signed rank tests. * p < 0.05, ** p < 0.01, *** p < 0.001, and n.s., not significant.

**Figure S2.**
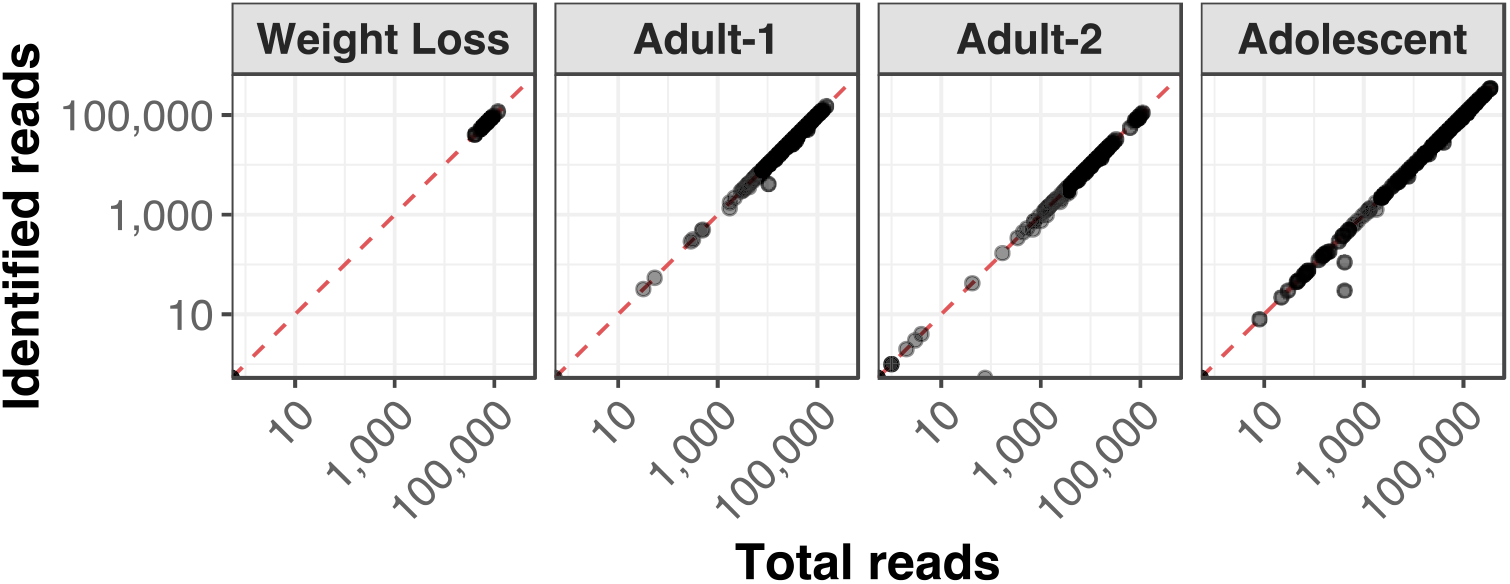
Total and identified sequence reads by sample and study. Sequence reads were assigned to a plant taxon by exact matching to sequences in the custom *trnL* reference (see **Methods**). The red dashed line indicates a complete mapping (*i*.*e*., all reads in a sample are identified by at least one sequence in the *trnL* reference and assigned to a plant taxon).

**Figure S3.**
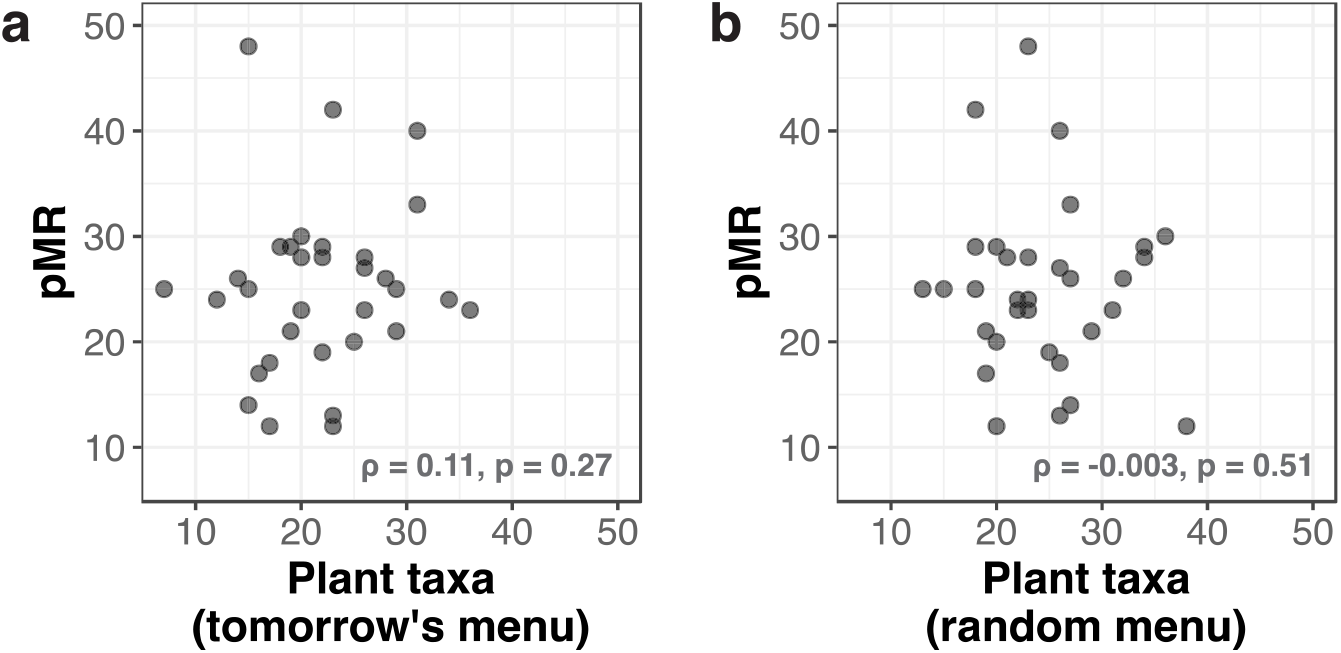
pMR is unrelated to menu data from days with no biological connection to tested stool samples in the Weight Loss cohort. One-tailed Spearman correlations between pMR and menu data from **(a)** the day after stool sample collection or **(b)** a random menu day.

**Figure S4.**
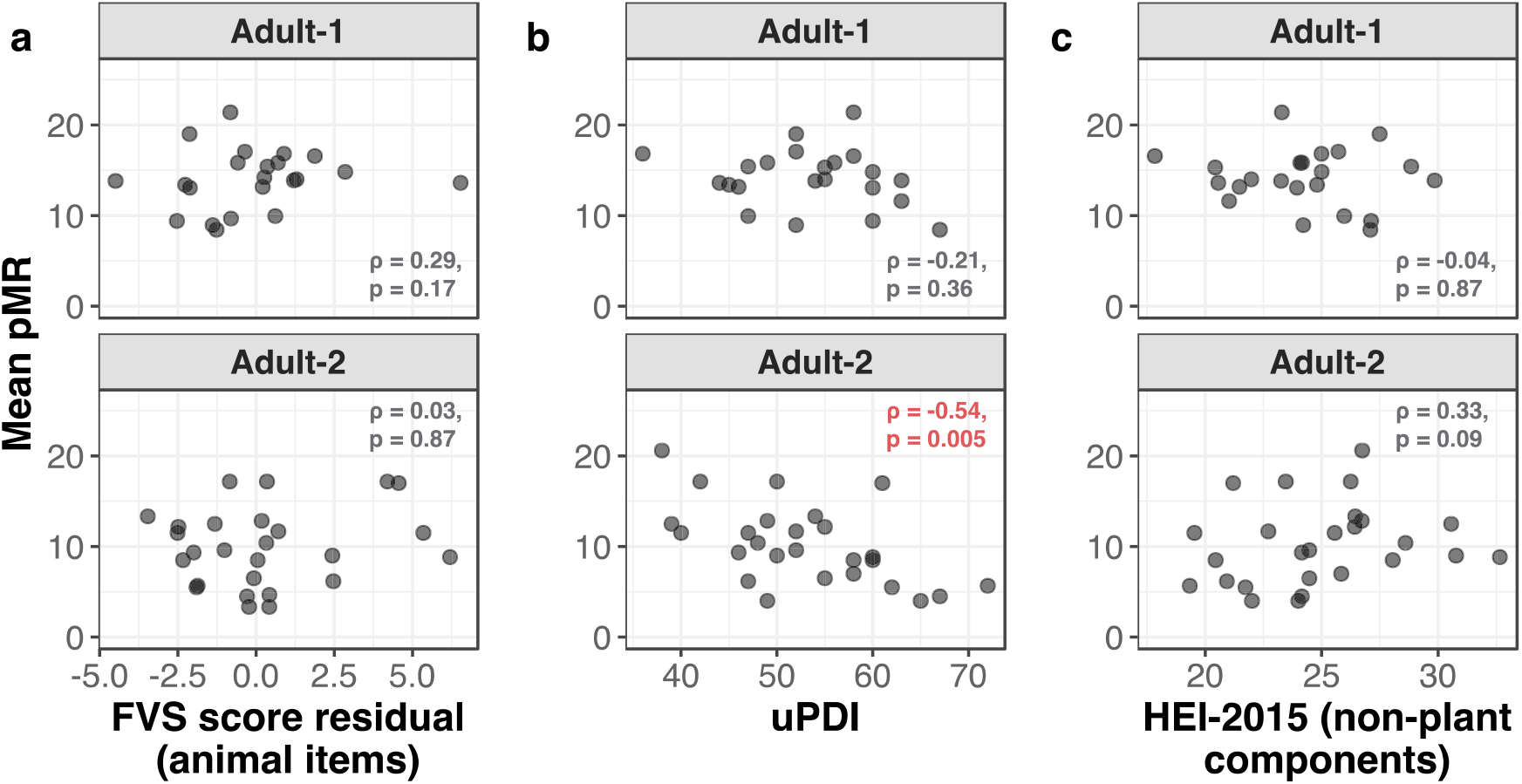
Correlations between pMR and animal-based or unhealthy plant component scores are absent or negative in the Adult-1 and Adult-2 cohorts. Correlations between mean pMR (pMR averaged across all available stool samples per participant) and animal- or unhealthy plant-based dietary diversity **(b)** and quality **(c, d)** indices derived from FFQ data in Adult-1 and Adult-2 participants.

**Figure S5.**
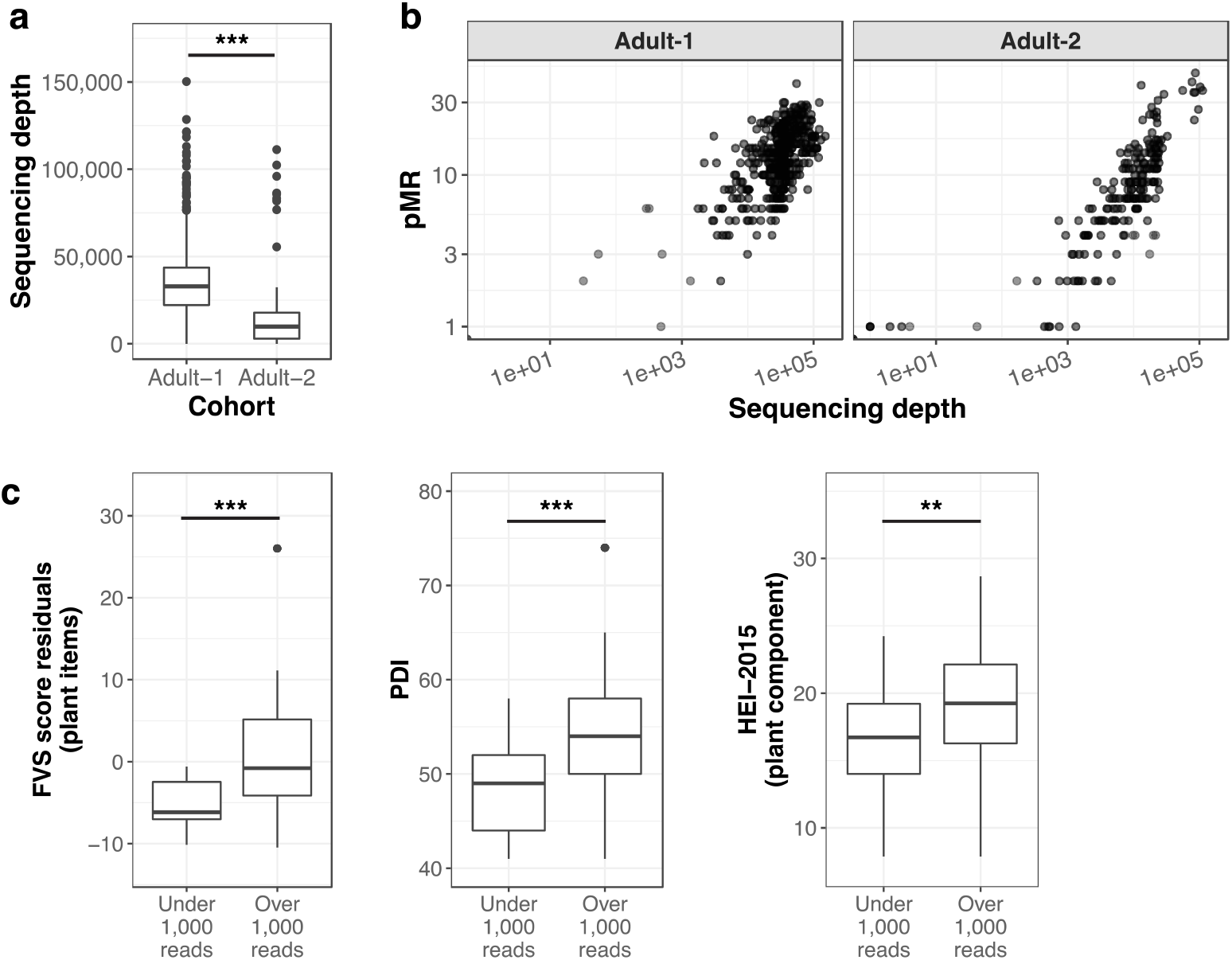
pMR scales with sequencing depth, which is influenced by biological factors. **(a)** *trnL* sequencing depth in Adult-1 and Adult-2 cohort samples. **(b)** Relationship between pMR and sequencing depth. **(c)** Survey-based measures of consumed plant diversity or plant-based diet quality by *trnL* sequencing depth. Statistical tests in **(a, c)** by Mann-Whitney U. ** p < 0.01, *** p < 0.001.

**Figure S6.**
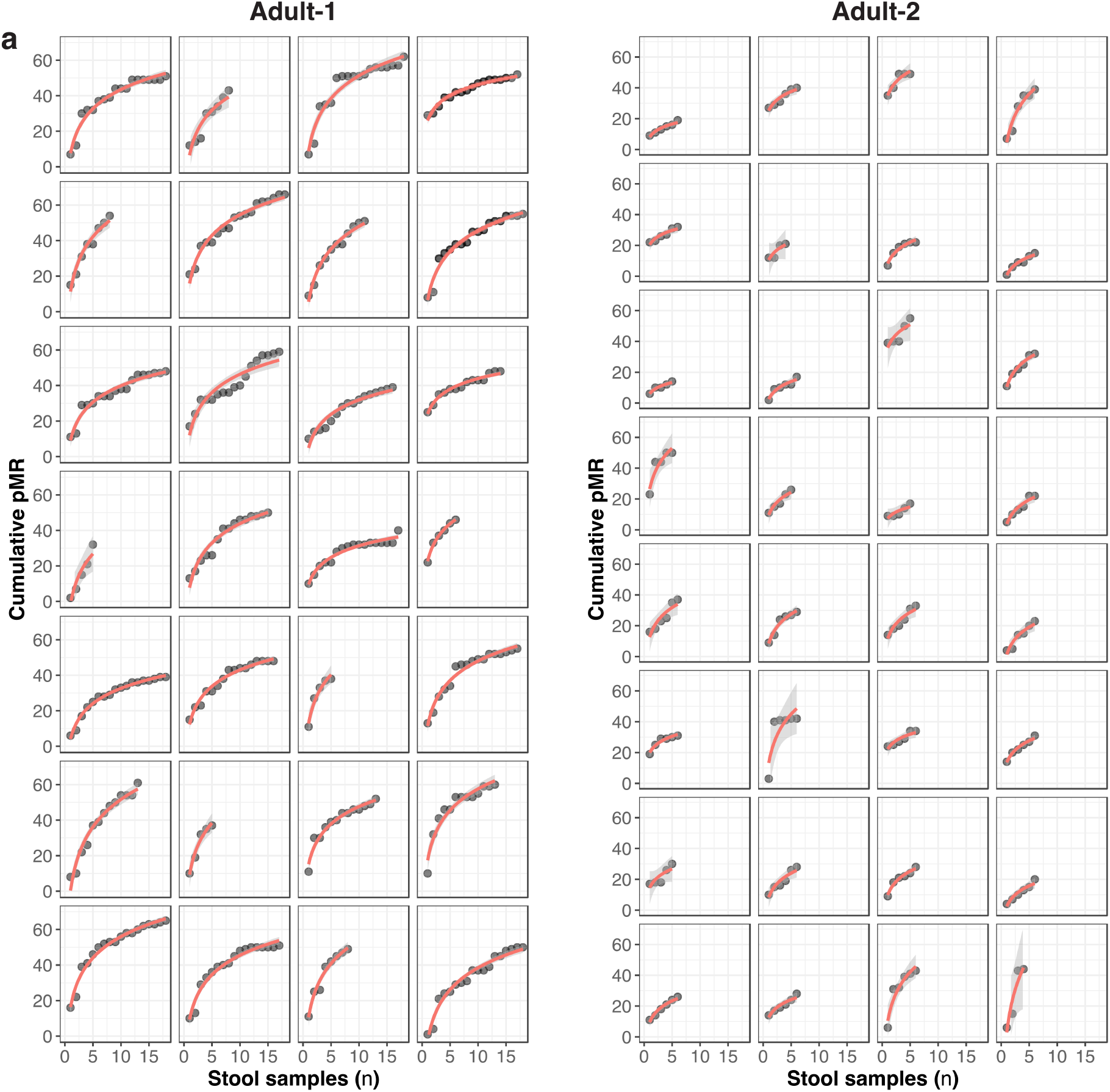
Sampling of Adult-1 and Adult-2 participants affects cumulative pMR. Cumulative pMR (# of unique *trnL* taxa observed across all samples) was calculated from each participant’s samples in consecutive order. Each facet represents one participant. Overlaid red curves are fits for the regression of cumulative pMR on log(sample number). Adult-1 participants collected at most 18 samples, and Adult-2 participants at most 6.

**Figure S7.**
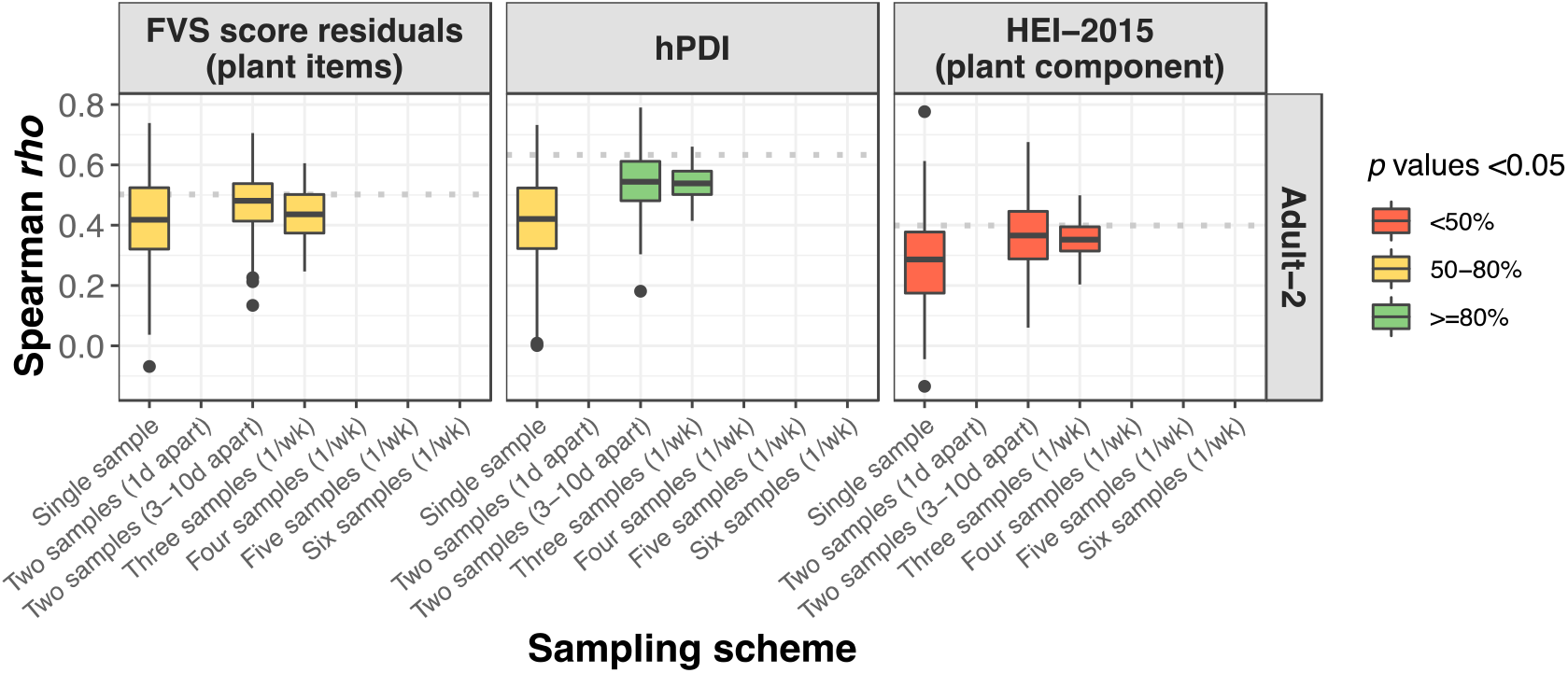
Subsampling of pMR-dietary index relationships in Adult-2 participants. Correlations from lower panels of **(Fig. 2b-d)** re-tested under candidate sampling schemes with mean pMR derived from a smaller number of stool samples. All boxplots represent ∼100 random subsamples at each strategy, and color indicates the percentage of iterations reaching the statistical significance threshold of p<0.05. Results are from two-tailed Spearman correlations.

**Figure S8.**
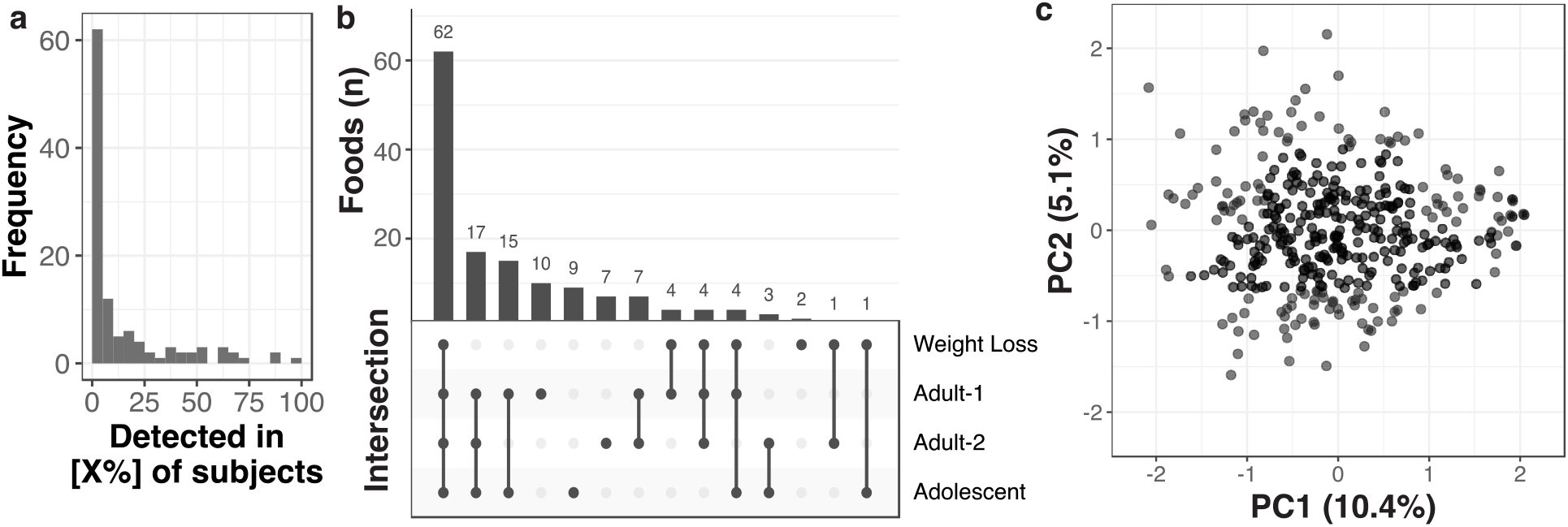
Dietary landscape of Adolescent participants. **(a)** Histogram of participant prevalence of plant taxa detected by *trnL* metabarcoding in the Adolescent cohort. Most foods were shared by a relatively small number of subjects (distribution skewness=1.84). **(b)** UpSet plot (an scalable alternative to Venn diagrams for visualizing intersecting sets^60^) indicating overlap of plant taxa detected by *trnL* metabarcoding in the Adolescent cohort and those detected in adult cohorts (Weight Loss, Adult-1, and Adult-2). The largest intersection is made up of plant foods detected across all four cohorts, and the second largest of plant foods detected in all free-eating cohorts. **(c)** Principal components analysis of presence-absence data of each *trnL* taxon in Adolescent samples. Each point indicates an individual sample projected onto the first two principal components, which capture 10.4% and 5.1% of the variance of the overall data, respectively.

**Table S1.** Baseline characteristics of trnL metabarcoding cohorts. All values are reported as mean ± standard deviation except samples per individual, which is given as median ± median absolute deviation. Entries for Adult-1 and Adult-2 race do not sum to 100% due to missing raw data (*i*.*e*. individuals that did not indicate a response).

|  | Weight Loss | Adult-1 | Adult-2 | Adolescent |
| --- | --- | --- | --- | --- |
| ***n*** |  |  |  |  |
| Individuals | 4 | 28 | 32 | 246 |
| Samples/individual | 11.5±2.2 | 16.0±3.0 | 6.0±0.0 | 2.0±0.0 |
| Total samples | 41 | 387 | 189 | 384 |
| **Diet** |  |  |  |  |
| Type | Interventional reduced calorie | Free-eating with fiber supplement | Free-eating with fiber or placebo snack bar | Free-eating |
| Assessment | Digital menu system | FFQ (NCI DHQ3) | FFQ (NCI DHQ3) | Custom survey |
| **Demographics** |  |  |  |  |
| Age, years | 58.5±8.8 | 33.3±12.0 | 25.6±5.2 | 13.3±2.3 |
| Sex, % female | 50 | 39 | 59 | 60 |
| Race, % |  |  |  |  |
| Black | 0 | 4 | 3 | 53 |
| White | 75 | 68 | 44 | 38 |
| Asian | 0 | 11 | 38 | 2 |
| American Indian/<br>Alaska Native | 25 | 0 | 0 | 0 |
| Multiple | 0 | 7 | 9 | 7 |
| Ethnicity, % Hispanic | 0 | 11 | 13 | 18 |
| **Health** |  |  |  |  |
| BMI | 35.5±4.6 | 24.7±2.4 | 22.9±2.3 | 31.8±10.1 |

**Table S2.**
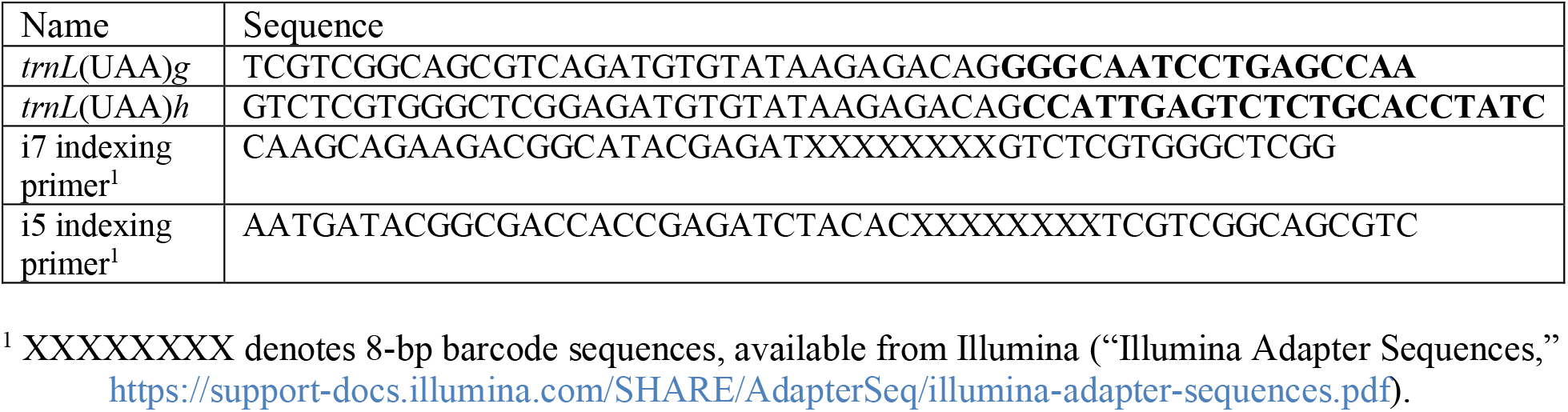
Primers used in this study. *trnL*-specific sequences are bolded and Illumina adapters are in standard typeface.

**Table S3.**
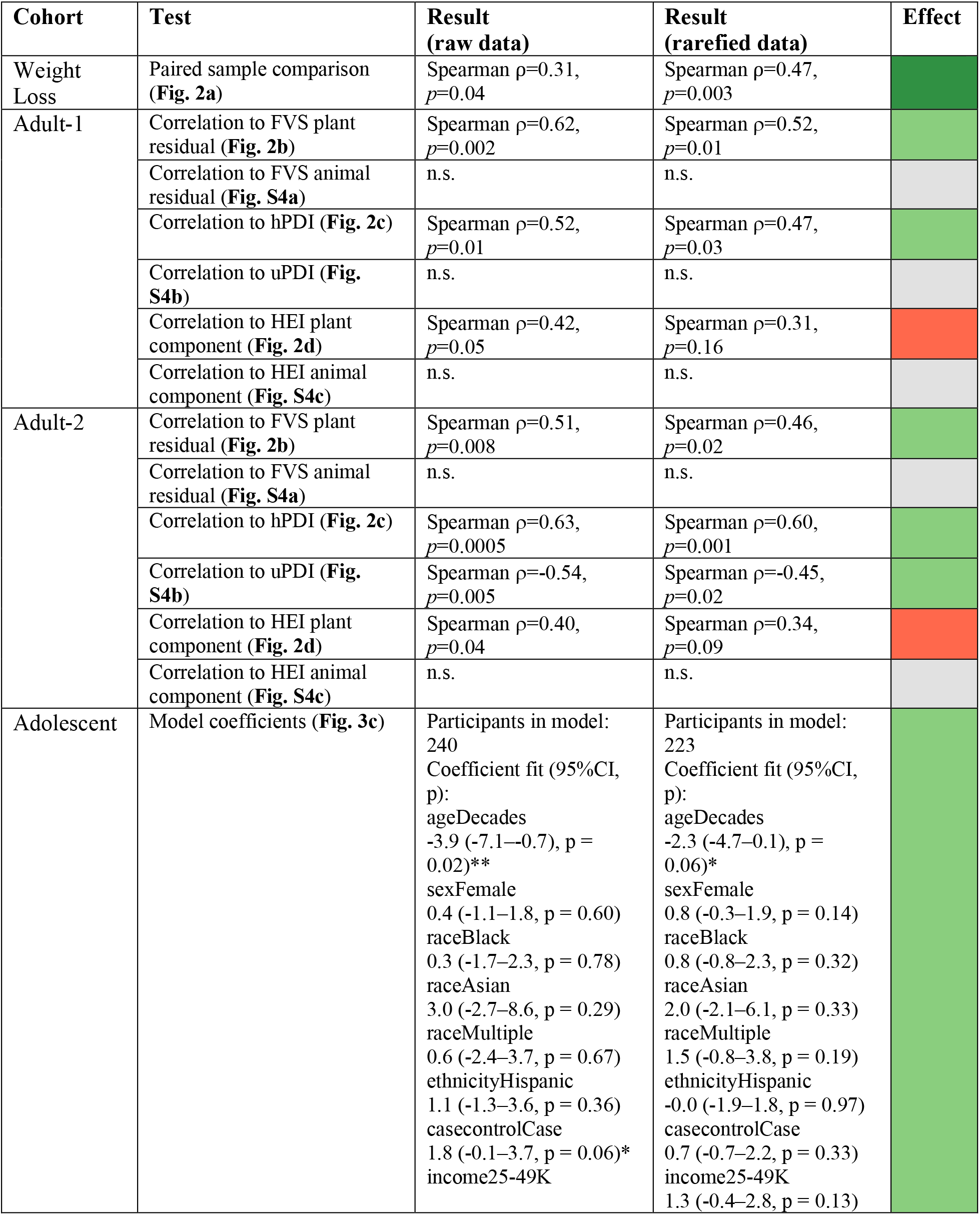
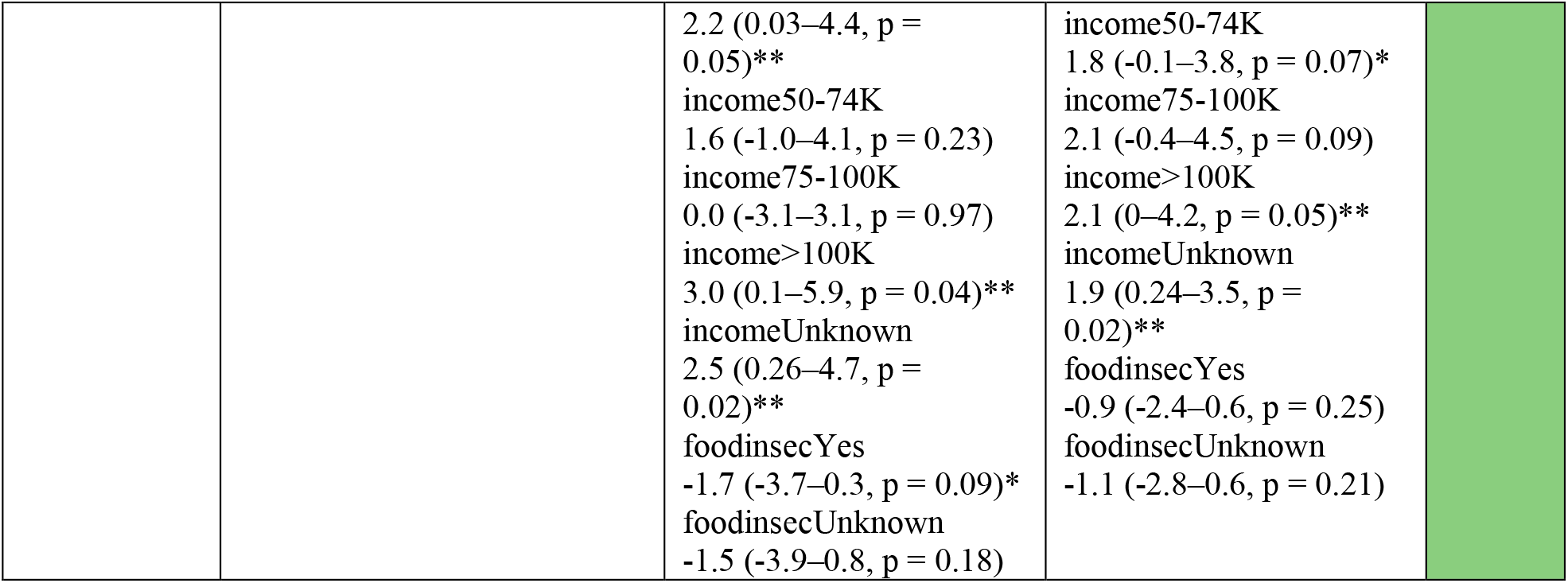
Impact of rarefaction on key study results. pMR was estimated at a shared read depth of 1,000 reads and any samples below that threshold excluded from the analysis. Color indicates impact of rarefaction: dark green, strengthened; light green, attenuated but still significant; red, not significant after rarefaction; gray, not significant and unchanged from raw result. Weight Loss participants, who ate diets designed to be healthful and high in plant foods, had no low read depth samples among those amplified, and thus rarefaction served exclusively to remove variation in pMR signal due to read depth, strengthening the detected relationship to menu data. In contrast, Adult-1, Adult-2, and Adolescent samples below the rarefaction threshold may convey meaningful information on low plant intake and attenuate relationships when excluded. * p < 0.10 (trend), ** p < 0.05.

